# Emergence of Genetic Mutations associated with Malaria Diagnostic and Artemisinin Partial Resistance in Somalia: A Genomic Surveillance Study

**DOI:** 10.64898/2026.07.19.26357122

**Authors:** Abdulkadir M. Arale, Abdikarim H Hassan, Abdinasir I. Mahmoud, Josh Rey, Irene Molina-de la Fuente, Ana Chopo-Pizarro, Tamara Yap, Abdullahi M. Hassen, Jamal Amran, Jane Cunningham, Marian Warsame, Khalid B Beshir

## Abstract

Histidine-rich protein 2 (HRP2)-based rapid diagnostic tests (RDTs) are central to malaria case management in Africa but fail when *Plasmodium falciparum* parasites lack the *pfhrp2* or *pfhrp3* genes. Widespread deletions have been reported in Eritrea, Ethiopia, and Djibouti, yet no systematic data have been available from Somalia. Between May and October 2023, we collected 7148 dried blood spot (DBS) samples from patients with suspected malaria attending eight health facilities across seven regions in Somalia. Field HRP2/pan-lactate dehydrogenase (LDH) RDTs and microscopy were performed, and DNA was extracted from 301 RDT-positive and 173 RDT-negative DBS samples. A multiplex quantitative PCR assay targeting *pfldh, pfhrp2*, and *pfhrp3* was used to identify deletions in *pfldh*-positive samples lacking *pfhrp2* or *pfhrp3* amplification, with mixed infections inferred from delta cycle threshold (ΔCt) differences.

Of 474 analysed samples, 301 (4.2%, 95% CI 3.7-4.7) were RDT or microscopy positive, and 159 (33.5%) were confirmed *pfldh*-positive by qPCR. Among these, six (3.8%, 95% CI 1.4-8.1) carried *pfhrp2* deletions and 59 (37.1%, 95% CI 29.6-45.1) carried *pfhrp3* deletions. Eleven infections (6.9%, 95% CI 3.5-12.1) produced discordant RDT outcomes, HRP-/LDH+ or RDT-negative despite *pfldh* positivity. Deletions were most frequent in Dolow, Luq, and Bosaso. A single isolate carried the *pfk13* R622I mutation, confirming the first report of the emergence of an artemisinin partial resistance-associated in Dolow, Gedo region, Somalia.

*Pfhrp2/3* deletions causing false RDT results remain low in Somalia and the confidence interval overlaps with the 5% policy threshold for changing RDTs, indicating uncertainty that warrants larger-scale assessment. *Pfhrp3* deletions are widespread and compromise the diagnostic redundancy of HRP2-based tests. Most deletion-carrying parasites remain detectable through the pan-LDH line, minimising immediate clinical risk but leading to systematic misclassification of *P. falciparum* as non-falciparum malaria. These findings support the continued use of HRP2/Pan-LDH RDTs but highlight high risk areas and emphasise the need for periodic and expanded molecular surveillance for prevalence trends to guide timely future diagnostic policy.

## Introduction

Rapid diagnostic tests (RDTs) based on detection of *Plasmodium falciparum* histidine-rich protein 2 (*Pf*HRP2) have transformed malaria case management across Africa. They offer a rapid, reliable alternative to microscopy in resource-limited settings, where laboratory capacity is often minimal. However, deletions in the *pfhrp2* gene and its paralogue *pfhrp3* undermine test performance by preventing HRP2 antigen expression, resulting in false-negative results, misdiagnosis, and under-reporting of *P. falciparum* infections(1). This diagnostic escape poses a direct threat to both individual patient care and national surveillance systems, jeopardising malaria elimination efforts(2).

The Horn of Africa has become the epicentre of *pfhrp2/3* deletions. Eritrea was the first country to report widespread deletions causing false negative RDTs exceeding WHO’s 5% threshold for diagnostic policy change, leading to the replacement of HRP2-only RDTs with LDH-based formats(3). Ethiopia has documented substantial heterogeneity, with *pfhrp3* deletions widespread and *pfhrp2* deletions emerging in multiple regions(4, 5). Djibouti has recorded high false-negative rates attributable to these deletions (6), while Sudan has also con(7)firmed *pfhrp2*-negative parasites(8). Beyond the Horn, deletions have been detected in South Sudan, Kenya, Tanzania, Ghana, and the Democratic Republic of Congo(1, 9–12), suggesting that this phenomenon is emerging and/or spreading across the continent.

The implications of *pfhrp2/3* deletions differ by national treatment protocol. Somalia follows a unified case management policy: all confirmed malaria infections are treated with artemisinin-based combination therapy (ACT) regardless of species, reflecting the predominance of *P. falciparum* and the limited role of *P. vivax(13)*. In contrast, Ethiopia retains a species-specific regimen, ACT for *P. falciparum* and chloroquine ± primaquine for *P. vivax*. Misclassification in Ethiopia therefore risks chloroquine treatment failure for undetected *P. falciparum* infections. In Somalia, the risk is reversed: *pfhrp2*-deleted *P. falciparum* infections producing false-negative or pan-only RDT results may be recorded as non-falciparum cases or missed entirely, leading to diagnostic uncertainty and potential under-treatment in peripheral facilities, where species confirmation by microscopy may not be available(14).

Concurrently, the emergence of artemisinin resistance threatens malaria control on a different front. Mutations in the *P. falciparum* kelch13 (*pfk13*) gene, first associated with delayed parasite clearance in Southeast Asia, have now been detected across East Africa, The R561H mutation has expanded clonally in Rwanda and Tanzania, while A675V and C469Y are prevalent in northern Uganda(15–18). In Eritrea and Ethiopia, the R622I mutation has recently emerged in parallel with high *pfhrp2/3* deletion prevalence, suggesting concurrent selection pressures on diagnostic and drug resistance loci(4, 15, 16, 19). The co-existence of these threats in neighbouring countries signals an evolving epidemiological convergence that could undermine both treatment and diagnosis(4, 15).

Somalia remains highly malaria-endemic, with *P. falciparum* responsible for most infections and *P. vivax* present at lower levels. Transmission is heterogeneous and seasonal, concentrated along riverine and coastal areas. The health system relies almost entirely on HRP2-based RDTs for diagnosis; any compromise in their reliability could have severe consequences for case management and reporting accuracy. Yet, despite its geographical proximity to Eritrea, Ethiopia, and Djibouti, Somalia has lacked systematic data on *pfhrp2/3* deletions or *pfk13* mutations. This study aimed to fill the existing data gap by conducting multisite molecular surveillance across southern and central Northeast Somalia to determine the prevalence and distribution of *pfhrp2/3* deletions and *pfk13* polymorphisms, evaluate their impact on RDT performance, and assess implications for national diagnostic and treatment policy.

## Methods

### Study sites and design

This cross-sectional molecular surveillance was conducted between May and October 2023 to assess *P. falciparum* parasites for *pfhrp2/3* gene deletions and *pfk13* mutations. Eight health facilities were selected in consultation with the National Malaria Control Programme to represent the country’s main epidemiological zones: Wadajir (Banadir region), Bosaso (Bari), Baidoa (Bay), Dolow and Luq (Gedo), Beledweyne (Hiran), Afgoi (Lower Shabelle), and Balad (Middle Shabelle). The selection reflected both transmission intensity and logistical feasibility within the surveillance network.

### Participant recruitment and field data collection

Patients of all ages presenting with fever or a history of fever at the selected facilities were screened for malaria as part of routine case management. Finger-prick blood samples were used for HRP2/pLDH rapid diagnostic testing (Bioline™ Malaria Ag P.f/Pan, Abbott Diagnostics Korea-05FK60) and preparation of dried blood spots (DBS) on Whatman filter paper. RDT results were interpreted by trained health workers according to the manufacturer’s instructions. Thick and thin blood films were prepared and read by trained microscopists at the study health facilities for cross-validation of RDT results. The blood slides were stained with 2.5% Giemsa to detect asexual malaria parasites and determine parasite species. A slide was considered negative when examination of 1000 white blood cells reveal no asexual parasites. DBS samples were dried, stored with desiccants, and transported under cold chain to the central reference laboratory at the London School of Hygiene & Tropical Medicine (LSHTM) for molecular analysis.

DNA was extracted from DBS samples using robotic DNA extraction following manufacturer’s instructions (Qiagen, Germany) using published protocol (20). Extraction quality was verified through amplification of the human β-tubulin gene as an internal control. Parasite DNA was screened using a validated multiplex quantitative PCR (qPCR) assay targeting *pfldh* (parasite control gene) together with *pfhrp2* and *pfhrp3(21)*. Samples with *pfldh* cycle threshold (Ct) values greater than 35 were excluded from analysis to avoid stochastic amplification at low parasite densities. Deletions were defined as the absence of *pfhrp2* or *pfhrp3* amplification in the presence of a valid *pfldh* signal. Mixed infections were identified when the difference between *pfhrp2* or *pfhrp3* and *pfldh* Ct values (ΔCt) exceeded 2·5 cycles, consistent with previously published criteria(21).

Detection of *pfk13* mutations associated with artemisinin resistance was carried out by amplicon deep sequencing using Oxford Nanopore Technologies (ONT). The *pfk13* propeller domain was amplified by PCR from all *pfldh*-positive samples and prepared for sequencing using the ONT Native Barcoding Kit (SQK-NBD114-96)(15). Barcoded libraries representing up to 96 samples per run were pooled, end-prepped, and sequenced on MinION flow cells (R10.4.1). Basecalling and demultiplexing were performed using Guppy v6.5.7 (ONT, UK). Reads were filtered by Q-score ≥ 9, trimmed, and aligned to the *P. falciparum* 3D7 reference genome (PF3D7_1343700) using minimap2. Variant calling was conducted with Medaka and bcftools, and amino acid substitutions were annotated against WHO-validated and candidate resistance markers (22) using R script.

Field and laboratory results were entered into a central database and cross-checked for consistency. Statistical analyses were performed in R version 4·3·2 (R Foundation, Austria). The prevalence of *pfhrp2/3* deletions was calculated as the proportion of *pfldh*-positive samples lacking amplification of either or both target genes. Proportions and 95% confidence intervals were computed by region and overall. Correlation between field RDT results and molecular deletion status was examined to identify discordant or false-negative outcomes.

This work formed part of routine national molecular surveillance supported by the World Health Organization and LSHTM and did not require formal ethics committee review, as no additional sampling or patient interventions were undertaken beyond standard malaria case management.

## Results

### Field diagnostics

Of the total 7148 samples, 301 (4.2%, 95% CI 3·7–4·7) were positive by either RDT or microscopy. Concordance between the two diagnostic methods was high overall, though 11 cases (3·6%, 95% CI 1·8–6·3) were discordant (microscopy positive/RDT negative), mainly showing HRP2–/LDH+ profiles suggestive of potential *pfhrp2/3* deletions or non-falciparum infections. RDT positivity varied markedly across districts, from 0·1% (95% CI 0·0–0·6) in Wadajir (Banadir) to 8·9% (95% CI 6·3–12·4) in Beledweyne (Hiran; Table 1)

**Table 1.** RDT and Microscopy results by region RDT and microscopy results by district in Somalia. The table shows the total number of samples tested, the number positive by RDT and microscopy, and the proportion of positives expressed as percentages. *Discordant cases (HRP–/LDH+ or RDT negative despite microscopy positive) are also noted. Overall, out of 7148 samples, 301 (4.2%) were RDT positive and 238 (3.3%) were microscopy positive, with 11 discordant results observed. Discordant outcomes were concentrated in Bosaso, Baidoa, Dolow, and Afgoi, highlighting locations where pfhrp2/3 deletions may compromise RDT performance.

| Region | District | Total screened | RDT POS (n, %) | Mic POS (n, %) | Comment |
| --- | --- | --- | --- | --- | --- |
| **Banadir** | Wadajir | 988 | 1 (0.1%) | 0 (0.0%) | No discordant* |
| **Bari** | Bosaso | 301 | 24 (8.0%) | 24 (8.0%) | 3 discordant |
| **Bay** | Baidoa | 1003 | 9 (0.9%) | 9 (0.9%) | 1 discordant |
| **Gedo** | Dolow | 1002 | 60 (6.0%) | 29 (2.9%) | 2 discordant |
|  | Luq | 998 | 51 (5.1%) | 51 (5.1%) | 0 discordant |
| **Hiran** | Beledweyne | 361 | 32 (8.9%) | 32 (8.9%) | No discordant |
| **L. Shebelle** | Afgoi | 993 | 35 (3.5%) | 40 (4.0%) | 5 discordant |
| **M. Shebelle** | Balad | 478 | 8 (1.7%) | 8 (1.7%) | No discordant |
| **Total** | — | **7148** | **301 (4.2%)** | **238 (3.3%)** | **11 discordant** |

### qPCR results

A total of 474 samples were included for molecular analysis, comprising all positives samples by microscopy or RDT (n=301) and a subset of randomly selected negatives from each region (n=173 total). Multiplex qPCR confirmed 159 of 474 samples (33·5%, 95% CI 29·4–38·0) *P. falciparum*-positive infections based on *pfldh* amplification. The proportion of *pfldh*-positive samples varied substantially by district, from 2·8% (95% CI 0·1–14·2) in Wadajir (Banadir) to 74·0% (95% CI 63·0– 83·0) in Afgoi (Lower Shabelle). Amplification success among *pfldh*-positive samples was 153 of 159 (96·2%, 95% CI 91·9–98·6) for *pfhrp2* and 101 of 159 (63·5%, 95% CI 55·5–71·0) for *pfhrp3* (Table 2). *Pfhrp3* amplification was markedly lower in Beledweyne, Luq, and Bosaso, consistent with a high prevalence of *pfhrp3* deletions in these districts.

**Table 2.** Multiplex qPCR results by District Summary of human DNA, pfldh, pfhrp2, and pfhrp3 positive results by district. The table shows the number of samples per district with successful human DNA amplification (quality control) and positive qPCR results for pfldh, and negative qPCR results for pfhrp2, and pfhrp3 targets. While all districts showed strong amplification of human DNA, positivity rates for pfldh varied. Pfhrp3 negativity was notably high in several districts, including Beledweyne, Bosaso, and Luq, consistent with widespread pfhrp3 deletions.

| District | hum DNA POS | *pfl dh* POS | *pfhrp2* NEG | *pfhrp3* NEG |
| --- | --- | --- | --- | --- |
| AFGOI | 50 | 37 (74.0%) | 0 (0.0%) | 0 (0.0%) |
| Baidoa | 42 | 10 (23.8%) | 1 (10.0%) | 3 (30.0%) |
| Balad | 38 | 7 (18.4%) | 0 (0.0%) | 0 (0.0%) |
| Beledweyne | 47 | 5 (10.6%) | 0 (0.0%) | 4 (80.0%) |
| Bosaso | 42 | 15 (35.7%) | 3 (20.0%) | 11 (73.3%) |
| Dolow | 98 | 47 (48.0%) | 2 (4.3%) | 27 (57.4%) |
| Luq | 110 | 38 (34.5%) | 0 (0.0%) | 14 (36.8%) |
| Wadajir | 36 | 1 (2.8%) | 0 (0.0%) | 0 (0.0%) |
| Total | 474 | 159 (33.5%) | 6 (3.8%) | 59 (36.5%) |

### *Pfhrp2/3* deletions

Among the 159 *pfldh*-positive samples, six (3·8%, 95% CI 1·4–8·1) *were pfhrp2/3 negative* and 59 (36.5%, 95% CI 29·6–45·1) were *pfhrp3* negative. No district recorded *pfhrp2* negative alone. The highest proportions of *pfhrp3*-negative infections were observed in Dolow (27/47, 57·4%, 95% CI 42·2–71·7), Luq (14/38, 36·8%, 95% CI 22·7–52·9), and Bosaso (11/15, 73·3%, 95% CI 44·9–92·2) (Table 2). By contrast, *pfhrp2/3* negatives were rare, detected sporadically in Dolow, Luq, and Bosaso.

Using the cycle threshold (Ct) values, we applied the criteria described in the Methods to determine whether *pfhrp2* and *pfhrp3* negatives represented true gene deletions. Scatter plots comparing *pfhrp2* and *pfhrp3* Ct values against *pfldh* (Figure 2) showed distinct amplification patterns across the sample set. Of the 159 *pfldh*-positive samples, 100 (64·2%) had Ct values below 35 for all three genes, indicating intact loci and no evidence of deletion (grey points). Fifty-nine samples (28·9%) showed delayed or absent *pfhrp2/3* or *pfhrp3* amplification with Ct > 35 while *pfldh* remained strongly positive, consistent with low-level or mixed infections containing both deleted (blue and red points) and non-deleted clones (grey points). Of the 59 samples, six (6/159, 3.8%) showed complete absence of *pfhrp2* and *pfhrp3* amplification despite robust *pfldh* signals, representing confirmed gene deletions (blue points)

**Figure 1.**
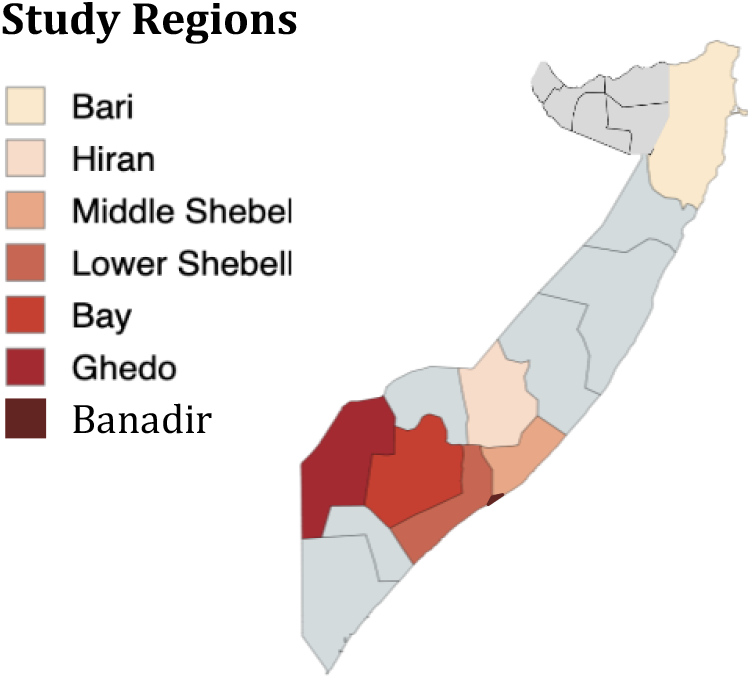
Map of Somalia showing study regions. Highlighted regions indicate where health facilities were included in the pfhrp2/3 surveillance study: Banadir (Wadajir), Bari (Bosaso), Bay (Baidoa), Gedo (Dolow and LUQ), Hiran (Beledweyne), Lower Shabelle (Afgoi), and Middle Shabelle (Balad). The remaining districts are shaded in grey and were not part of the study.

**Figure 2.**
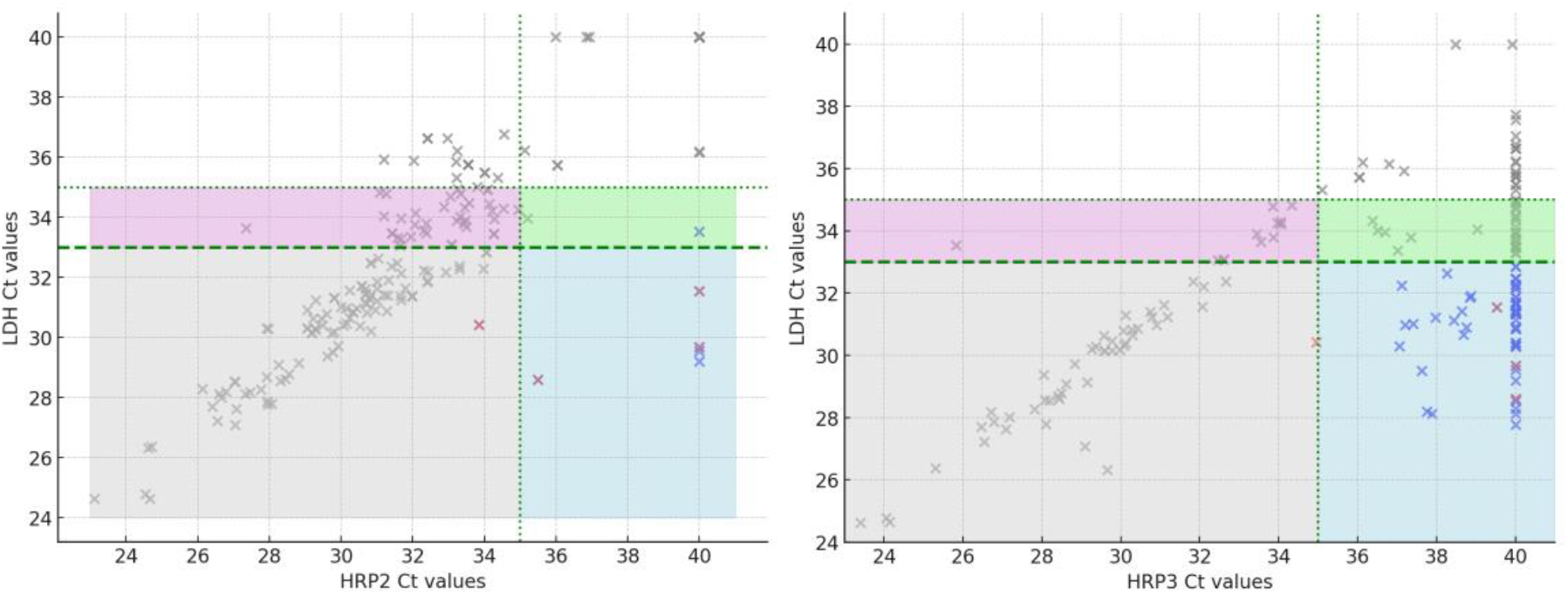
Scatterplots of P. falciparum HRP2 (left) and HRP3 (right) qPCR cycle threshold (Ct) values against LDH Ct values in Somalia samples. Each point represents an individual sample. The green dashed horizontal line marks the LDH Ct threshold (Ct = 33) for parasite positivity, while the vertical dotted line marks the HRP2 or HRP3 Ct threshold (Ct = 35). The shaded grey areas correspond to concordant positives for both HRP and LDH. The shaded pink and green areas represent borderline Ct values (33–35), consistent with low-density infections. The shaded blue area indicates samples with high HRP2/3 Ct (≥35) but positive LDH, consistent with pfhrp2 or pfhrp3 deletions. Red and blue points highlight samples with discordant results or confirmed deletions, respectively.

### Impact on RDT performance

Of the 11 discordant samples, five samples with either qPCR negative (n=4) or having low parasite density (n=1) were excluded from the analysis. Of the 159 molecularly confirmed *P. falciparum* infections, four (2·5%, 95% CI 0·7–6·3) were associated with discordant or false-negative HRP2-based RDT results (red points, Figure 2). Three of these showed HRP–/LDH+ profiles, and one was completely RDT-negative despite clear *pfldh* amplification. Most discordant samples originated from Dolow (bordering Ethiopia), Bosaso, and Baidoa districts that also showed the highest *pfhrp3* deletion prevalence. Two discordant samples were classified as *P. vivax* by microscopy, yet qPCR analysis revealed *P. falciparum* carrying *pfhrp2/3* deletions in mixed infections. Without molecular confirmation, these would have been recorded as *P. vivax-*only cases, underestimating the true *P. falciparum* burden. This highlights how *pfhrp2/3* deletions can bias routine surveillance data and obscure accurate species attribution in co-endemic settings.

Importantly, five of six (83·3%, 95% CI 43·7–97·0) discordant or RDT-HRP2-negative infections were still detected by the pan-LDH test line, mitigating the risk of untreated malaria but potentially causing misclassification as non-falciparum infections.

#### *Pfk13* mutations

Amplicon sequencing of the *pfk13* propeller domain was successfully completed for 147/159 (92·5%) *pfldh*-positive samples. One isolate from Dolow carried a nonsynonymous R622I mutation, corresponding to a validated marker of partial artemisinin resistance. The remaining sequences showed wild-type alleles at all other validated and candidate loci (including C469Y, A675V, and R561H). The observed R622I frequency was 0·7% (95% CI 0·02–3·9) of all *pfk13*-sequenced samples, representing the first confirmed detection of this mutation in Gedo (Dolow district).

## Discussion

This surveillance provides the first systematic evidence of *pfhrp2/3* deletions and *pfk13* polymorphisms in Somalia. The results show that *pfhrp2* deletions are present and *pfhrp3* deletions are considerably more widespread, mirroring the pattern observed in neighbouring Horn of Africa countries(4, 15). Although the findings suggest that the overall prevalence of deletions causing false-negative RDTs is currently low, the final sample size after qPCR confirmation was reduced from 301 to159 and confidence intervals cross 5%. More surveillance with larger sample sizes is needed to accurately determine the true probability that the country has surpassed the threshold for changing RDTs. Reassuring was the absence of dual deletion and that most *pfhrp2*-deleted samples were detected on pLDH test line. The identification of a single *pfk13* R622I mutation marks the first evidence of artemisinin-resistance–associated mutation in the Gedo region, bordering Ethiopia, underscoring the need for integrated surveillance of both diagnostic and therapeutic markers in the region.

The pattern observed in Somalia, frequent *pfhrp3* deletions and focal *pfhrp2* loss, resembles that of Eritrea and Ethiopia, where *pfhrp3* deletions are widespread and *pfhrp2* deletions are expanding under RDT selection pressure(4, 15). In Eritrea, deletion frequencies exceeding 40% prompted replacement of HRP2-only tests with LDH-based RDTs(3). Djibouti has similarly reported high false-negative rates linked to *pfhrp2/3* deletions(6), and Sudan has confirmed sporadic *pfhrp2*-negative isolates(8). This gradient suggests that *pfhrp3* loss often precedes *pfhrp2* deletion expansion. The persistence of *pfhrp3* deletions may reflect reduced diagnostic pressure, as HRP3 cross-reactivity with HRP2 antibodies partially preserves test positivity(1).

The clinical significance of *pfhrp2/3* deletions lies in their impact on HRP2-based RDTs, which remain the mainstay of malaria diagnosis in Somalia. Despite emerging deletions, clinical impact is currently limited: fewer than 3% of infections produced false-negative HRP2-based RDT results, and most retained detectable pan-LDH reactivity, ensuring patients were likely treated as malaria cases. However, this creates a diagnostic paradox in that deletions do not eliminate detection entirely but shift species interpretation, leading to *P. falciparum* being misclassified as non-falciparum malaria. This misclassification can distort surveillance data and underestimate *P. falciparum* prevalence in mixed or low-density infections. Two discordant cases in this study were misclassified as *P. vivax* by microscopy, yet qPCR confirmed mixed infections containing *pfhrp2/3*-deleted *P. falciparum* clones. Taken together, these findings foreshadow the diagnostic trajectory observed in Eritrea, where deletions expanded rapidly and forced a national diagnostic policy shift(3, 15).

As per the WHO surveillance protocol template, the survey team aimed to enrol 300 *P. falciparum* cases; however, qPCR results confirmed a high prevalence of false-positive results from HRP2-based RDTs and microscopy with only 159 out of 301 *P. falciparum* positive RDT and/or microscopy results confirmed by qPCR. This possibly reflects limited specificity of antigen- and morphology-based detection methods in low-transmission settings and/or operator or administrative errors. These false positive results can inflate surveillance data, distort test positivity rates, and lead to inappropriate treatment or misallocation of resources. Ensuring operators of RDTs and microscopy have appropriate training, supervision and periodic proficiency testing is essential to maintain reliable performance. qPCR provides a more accurate measure of active parasitaemia and should be used if available, periodically to recalibrate field diagnostics, particularly as Somalia approaches lower transmission thresholds.

Given the limited sensitivity of microscopy in detecting low-density infections, reliance on slide re-examination to resolve discordant RDT results may underestimate true *P. falciparum* prevalence. In such cases, pf-LDH–based RDTs offer a more practical and reliable alternative for identifying HRP2-negative but *P. falciparum*–positive infections, particularly in peripheral facilities with limited technical capacity. Adoption of dual-target RDTs that include pf-LDH detection would enhance diagnostic accuracy and provide a safeguard against HRP2-based test failure, especially as *pfhrp3* deletions continue to erode redundancy in HRP2-dependent assays.

The *pfk13* R622I mutation detected in one isolate from Dolow, near the Ethiopian border, and previously reported in Northeast warrants close monitoring. This substitution has been linked to delayed parasite clearance in Eritrea and reported sporadically in East Africa(4, 15, 23). Its emergence in Somalia likely reflects regional gene flow and underscores the permeability of cross-border transmission networks. No other validated or candidate *pfk13* mutations (C469Y, A675V, R561H) were found, consistent with the absence of clinical artemisinin resistance in Somalia(24).

Together, these findings suggest that Somalia remains in a pre-emergence phase for both diagnostic and drug resistance threats. HRP2-based RDTs remain reliable for routine case management, especially if accompanied by a pan-LDH test line that detects the majority of *pfhrp2*/*3* deleted parasites, as well as *P. vivax*, but early warning signals are evident. To preserve diagnostic integrity, the National Malaria Control Programme (NMCP) should maintain molecular monitoring of *pfhrp2/3* deletions and consider periodic evaluation of the performance of combination or LDH-based RDTs in sentinel sites. The NMCP should also monitor outcome of registration and WHO Prequalification processes for next generation RDTs which combined targets for HRP/pf-LDH on the same test line and *P. vivax* specific test lines which may circumvent entirely the challenges posed by *pfhrp2/3* deletions.

Several strengths of this study merit emphasis. The survey included multiple sites across southern and central Somalia, providing geographic breadth. The use of a validated multiplex qPCR assay ensured robust detection of deletions, with criteria for mixed infections applied consistently(21). Integration of field and laboratory data enabled identification of discordant cases and their likely molecular cause. The study also has limitations. Sample sizes in some districts were modest, limiting precision of prevalence estimates. As with all DBS-based analyses, low-density infections may yield false-negative qPCR results, although stringent Ct thresholds were applied to mitigate this. Finally, this was a cross-sectional snapshot and may not capture temporal changes in deletion prevalence.

Despite these limitations, the findings have programmatic implications. Routine molecular surveillance of *pfhrp2/3* deletions should be institutionalised within Somalia’s malaria monitoring framework, enabling early detection of shifts in prevalence. This should be coupled with strengthening of the quality of RDT and microscopy services. Diagnostic procurement policy should be reviewed, with consideration given to costs of ongoing surveillance versus phased adoption of anticipated next generation RDT combination tests.

The findings suggest *pfhrp2* deletions are not common in Somalia, but *pfhrp3* deletions are widespread and will already compromise the redundancy of HRP2-based diagnosis. The immediate clinical risk is limited by pan-LDH detection, but systematic misclassification of falciparum as non-falciparum malaria is occurring. The broader implication is that biological resistance, whether to diagnosis or treatment, rarely evolves in isolation. Somalia’s proximity to Eritrea, Djibouti and Ethiopia, its population mobility, and its dependence on a single diagnostic antigen all increase vulnerability to the spread of resistant parasite lineages. Sustained investment in genomic surveillance, strengthening in-country molecular capacity and linking laboratory data to case management and surveillance systems will ensure timely policy decisions. Periodic surveillance and adaptive diagnostic policies will be essential to prevent the trajectory observed in Eritrea and Djibouti, where *pfhrp2/3* deletions reached extremely high levels prior to changes to alternative RDTs.

## Data Availability

All data produced in the present study are available upon reasonable request to the authors

## References

1. Beshir KB, Sepulveda N, Bharmal J, Robinson A, Mwanguzi J, Busula AO, et al. Plasmodium falciparum parasites with histidine-rich protein 2 (pfhrp2) and pfhrp3 gene deletions in two endemic regions of Kenya. Sci Rep. 2017;7(1):14718.

2. World Health Organization. Response plan to pfhrp2 gene deletions, second edition. 2024.

3. Berhane A, Anderson K, Mihreteab S, Gresty K, Rogier E, Mohamed S, et al. Major Threat to Malaria Control Programs by Plasmodium falciparum Lacking Histidine-Rich Protein 2, Eritrea. Emerg Infect Dis. 2018;24(3):462–70.

4. Fola AA, Feleke SM, Mohammed H, Brhane BG, Hennelly CM, Assefa A, et al. Plasmodium falciparum resistant to artemisinin and diagnostics have emerged in Ethiopia. Nat Microbiol. 2023;8(10):1911–9.

5. Golassa L, Messele A, Amambua-Ngwa A, Swedberg G. High prevalence and extended deletions in Plasmodium falciparum hrp2/3 genomic loci in Ethiopia. PLoS One. 2020;15(11):e0241807.

6. Rogier E, McCaffery JN, Mohamed MA, Herman C, Nace D, Daniels R, et al. Plasmodium falciparum pfhrp2 and pfhrp3 Gene Deletions and Relatedness to Other Global Isolates, Djibouti, 2019-2020. Emerg Infect Dis. 2022;28(10):2043–50.

7. Lynch E, Jensen TO, Assao B, Chihana M, Turuho T, Nyehangane D, et al. Evaluation of HRP2 and pLDH-based rapid diagnostic tests for malaria and prevalence of pfhrp2/3 deletions in Aweil, South Sudan. Malar J. 2022;21(1):261.

8. Boush MA, Djibrine MA, Mussa A, Talib M, Maki A, Mohammed A, et al. Plasmodium falciparum isolate with histidine-rich protein 2 gene deletion from Nyala City, Western Sudan. Sci Rep. 2020;10(1):12822.

9. Bredu DG, Asamoah A, Adu GA, Abban BC, Anang SF, Peprah NY, et al. Prevalence of Plasmodium falciparum parasites with pfhrp2 exon 2 gene deletion in symptomatic malaria patients across Ghana in 2021. Malar J. 2025;24(1):170.

10. Kaaya RD, Kavishe RA, Tenu FF, Matowo JJ, Mosha FW, Drakeley C, et al. Deletions of the Plasmodium falciparum histidine-rich protein 2/3 genes are common in field isolates from north-eastern Tanzania. Sci Rep. 2022;12(1):5802.

11. Watson OJ, Slater HC, Verity R, Parr JB, Mwandagalirwa MK, Tshefu A, et al. Modelling the drivers of the spread of Plasmodium falciparum hrp2 gene deletions in sub-Saharan Africa. Elife. 2017;6.

12. Thomson R, Beshir KB, Cunningham J, Baiden F, Bharmal J, Bruxvoort KJ, et al. pfhrp2 and pfhrp3 Gene Deletions That Affect Malaria Rapid Diagnostic Tests for Plasmodium falciparum: Analysis of Archived Blood Samples From 3 African Countries. J Infect Dis. 2019;220(9):1444–52.

13. WHO. World Malaria Report. 2025.

14. World Heath Organisation. WHO guidelines for malaria. 2024.

15. Mihreteab S, Anderson K, Fuente IM, Sutherland CJ, Smith D, Cunningham J, et al. The spread of molecular markers of artemisinin partial resistance and diagnostic evasion in Eritrea: a retrospective molecular epidemiology study. Lancet Microbe. 2025;6(2):100930.

16. Mihreteab S, Platon L, Berhane A, Stokes BH, Warsame M, Campagne P, et al. Increasing Prevalence of Artemisinin-Resistant HRP2-Negative Malaria in Eritrea. N Engl J Med. 2023;389(13):1191–202.

17. Uwimana A, Legrand E, Stokes BH, Ndikumana JM, Warsame M, Umulisa N, et al. Emergence and clonal expansion of in vitro artemisinin-resistant Plasmodium falciparum kelch13 R561H mutant parasites in Rwanda. Nat Med. 2020;26(10):1602–8.

18. Ogwang R, Osoti V, Wamae K, Ndwiga L, Muteru K, Ningwa A, et al. A retrospective analysis of P. falciparum drug resistance markers detects an early (2016/17) high prevalence of the k13 C469Y mutation in asymptomatic infections in Northern Uganda. Antimicrob Agents Chemother. 2024;68(9):e0157623.

19. Zeleke AJ, Fola AA, Tollefson GA, Niare K, Leonetti A, Taropawala O, et al. Artemisinin resistant kelch13 R622I and RDT negativity approaching predominance in northern Ethiopia and emerging C580Y of African origin threaten falciparum malaria control. medRxiv. 2025.

20. Robinson A, Busula AO, Muwanguzi JK, Powers SJ, Masiga DK, Bousema T, et al. Molecular quantification of Plasmodium parasite density from the blood retained in used RDTs. Sci Rep. 2019;9(1):5107.

21. Grignard L, Nolder D, Sepulveda N, Berhane A, Mihreteab S, Kaaya R, et al. A novel multiplex qPCR assay for detection of Plasmodium falciparum with histidine-rich protein 2 and 3 (pfhrp2 and pfhrp3) deletions in polyclonal infections. EBioMedicine. 2020;55:102757.

22. WHO. Compendium of molecular markers for antimalarial drug resistance. 2025.

23. Menard D, Mihreteab S, Fidock DA. Artemisinin-Resistant HRP2-Negative Malaria in Eritrea. Reply. N Engl J Med. 2023;389(26):2497–8.

24. Warsame M, Hassan AM, Hassan AH, Jibril AM, Khim N, Arale AM, et al. High therapeutic efficacy of artemether-lumefantrine and dihydroartemisinin-piperaquine for the treatment of uncomplicated falciparum malaria in Somalia. Malar J. 2019;18(1):231.

